# Bridging the “Ten Walls” of Japanese Healthcare Data: A Comprehensive Semantic Mapping of JIPAD to HL7 FHIR R4 and Institutional Gap Analysis for the Japanese Health Data Space (JHDS)

**DOI:** 10.64898/2026.08.06.26359847

**Authors:** Kunihisa Ohno, Satoru Hashimoto

**Author notes:** Ethics Statement: This study did not involve patient data or personal information. All analyses were conducted using publicly available documentation (JIPAD Data Dictionary v3.7.2; FHIR R4 specifications; LOINC; SNOMED CT) and a non-identifying expert interview. Formal IRB review was not required.

## Abstract

**Background:** Japan faces critical challenges in medical data interoperability, conceptualized as the “Ten Walls” obstructing the Japanese Health Data Space (JHDS) [1]. The Japanese Intensive Care Patient Database (JIPAD) — Japan’s largest national ICU registry with 151 participating facilities — represents a high-quality critical care dataset that remains isolated from international data ecosystems.

**Objective:** To develop a formal mapping of all 122 JIPAD variables to HL7 FHIR R4, characterize the nature and magnitude of semantic gaps, and assess the feasibility of JIPAD integration into the JHDS.

**Methods:** All 122 JIPAD variables (Data Dictionary v3.7.2; Linkage Items List 20231020) were evaluated using ISO 21564 [8]-based semantic equivalence scoring across three tiers: High (direct FHIR R4 Core mapping), Partial (mapping via JP-Core Implementation Guide extensions [3]), and Low/No Equivalence (structural institutional gap). Semantically identical multi-instance fields (e.g., secondary disease codes ×5) were consolidated into single mapping entries, yielding 114 mapping entries. Pseudonymization architecture was characterized from primary documentation.

**Results:** Of 114 mapping entries representing the 122 JIPAD variables, 97 (85.1%) achieved High Equivalence via LOINC/SNOMED CT, and 12 (10.5%) achieved Partial Equivalence via JP-Core extensions, value-set translation, or FHIR R4 Core extension mechanisms — yielding a combined technical feasibility of 95.6% (109/114). Only 5 entries (4.4%) were classified as Low/No Equivalence, all attributable to Japan’s proprietary disease classification system (288 adult codes; 165 pediatric codes) embedded in the DPC reimbursement framework, plus one Japan-specific procedure (PMX endotoxin adsorption) absent from international terminology systems. Variable-level mapping details are provided in Supplementary Table S1. Critically, JIPAD employs pseudonymization with record-linkage capability, enabling 99% DPC data matching — demonstrating that technical and design-level barriers to FHIR integration have already been resolved.

**Conclusion:** JIPAD is technically and architecturally ready for FHIR integration at a 95.6% level. The remaining 4.4% barrier is exclusively institutional — rooted in MHLW policy frameworks governing the DPC disease classification system [6] — rather than technical. FHIR integration would further unlock pharmacoepidemiological and social epidemiological research currently inaccessible due to data isolation. As the sole national ICU registry providing high-acuity anchor data unavailable in general health records, JIPAD integration is essential for a clinically meaningful JHDS by 2027.

## 1. Introduction

The global convergence toward federated health data infrastructure — exemplified by the European Health Data Space (EHDS) [13] — demands that national clinical registries adopt internationally recognized semantic standards. Japan, however, faces a constellation of systemic barriers to this transition. Yamamoto (2026) has articulated one influential conceptual lens for these barriers, the “Ten Walls” framework [1], which we adopt here as a vocabulary for describing structural fragmentation of the Japanese Health Data Space (JHDS); the empirical analysis presented in this paper is independent of, and does not require commitment to, that specific typology.

The Japanese Intensive Care Patient Database (JIPAD), operated by the Japanese Society of Intensive Care Medicine (JSICM) under the ICU Function Evaluation Committee, represents Japan’s largest national ICU registry. As of 2025, JIPAD encompasses 151 facilities with over 200 registered cases each — qualifying those facilities for extended ICU reimbursement periods under Japan’s diagnosis-procedure combination (DPC) system. No other national Japanese registry captures comparable breadth of ICU-specific variables across this number of facilities; condition-specific registries such as J-SIPHE (infection surveillance) and ECMOnet (ECMO cases) address narrower clinical domains. JIPAD collects 122 standardized variables spanning patient demographics, severity scores (APACHE II/III, SOFA, PIM3), interventions, and outcomes.

Despite this clinical richness, JIPAD remains an isolated data island. Its integration into the JHDS, and by extension the emerging global health data ecosystem [14], has not been achieved. The purpose of this study is to determine whether this isolation reflects technical limitations, design-level constraints, or institutional policy barriers — and to provide empirical evidence for policymakers accordingly.

We present, to our knowledge, the first comprehensive semantic mapping of all JIPAD variables to HL7 FHIR R4, characterizing the nature, magnitude, and policy implications of each gap.

## 2. Methods

### 2.1 Data Sources

We utilized three primary documents as mapping substrates: (1) JIPAD Data Dictionary Version 3.7.2 (September 30, 2025), issued by the JSICM ICU Function Evaluation Committee; (2) JIPAD Linkage Items List (October 20, 2023), specifying all 122 data transmission variables with their CSV column assignments, nullability rules, and data types; and (3) JIPAD Disease Code List, cataloguing the proprietary disease classification system across 288 adult codes and 165 pediatric codes.

To characterize non-technical barriers to JIPAD–JHDS integration, one semi-structured interview was conducted with the JIPAD founding chair (S. Hashimoto) in March 2025. The interview was non-identifying and thematic, focusing exclusively on operational and institutional aspects of JIPAD; no patient data, personal data, or facility-identifying information was discussed. Themes addressed included: organizational capacity for FHIR implementation, communication channels with JHDS stakeholders, and the policy priority assigned to JIPAD integration within MHLW. Interview content was used solely to characterize the three non-technical barriers presented in Section 4.1 (organizational, epistemic, and administrative) and was not used to derive any of the variable-level mappings reported in Section 3. As the interview did not involve patient data and was conducted with a co-author in his expert capacity, formal IRB review was not required.

### 2.2 Mapping Framework

We employed a three-layer mapping framework targeting HL7 FHIR Release 4 (R4), which constitutes the current baseline specification for Japan’s national FHIR infrastructure and the JP-Core Implementation Guide; although FHIR R5 was published in 2023, R4 remains the operative standard for JHDS implementation planning as of 2025. First, Resource Identification assigned each variable to its corresponding FHIR R4 resource class (Patient, Encounter, Observation, Condition, Procedure, or MedicationAdministration). Second, Terminology Binding mapped each variable to LOINC or SNOMED CT codes where applicable. Third, Equivalence Assessment rated each mapping according to ISO 21564 [8] semantic equivalence criteria:

- High Equivalence: Direct semantic match within FHIR R4 Core [11] using paths and data types available without modification, with an established LOINC [9] or SNOMED CT [10] code where coded values are required. Profile-level conformance to JP-Core (e.g., JP-Core Patient profile constraints on FHIR R4 Core resources) does not, by itself, lower the assignment from High, provided no JP-Core extension is invoked.
- Partial Equivalence: Semantic match achievable without modification to FHIR R4 Core itself, via one of four mechanisms: (a) JP-Core Implementation Guide v1.1.1 extensions [3]; (b) value-set translation between JIPAD coded values and FHIR-conformant ValueSets (e.g., HL7 admit-source, discharge-disposition); (c) FHIR R4 Core extension mechanisms (e.g., Procedure.extension, Encounter.extension); or (d) use of existing FHIR R4 Core paths and resource structures (e.g., Procedure.performedPeriod, resource splitting into multiple Condition instances). Mechanism type for each Partial mapping is annotated in Supplementary Table S1.
- Low/No Equivalence: Structural gap arising from Japan-specific regulatory or administrative frameworks, for which no international semantic counterpart exists.

### 2.3 Equivalence Adjudication Protocol

To ensure reproducibility of equivalence assignments, each variable was adjudicated through a fixed decision sequence applied uniformly across the dataset. The sequence proceeds in four stages: (i) FHIR R4 Core resource identification; (ii) terminology binding evaluation against LOINC and SNOMED CT international releases; (iii) where no direct binding exists, evaluation of mapping mechanisms that preserve FHIR R4 Core unchanged — namely, JP-Core Implementation Guide v1.1.1 extensions, value-set translation, FHIR Core extension mechanisms, and use of Core paths or resource structures; and (iv) classification as institutional gap when no internationally-recognized semantic counterpart can be obtained through any of the preceding mechanisms. A variable was assigned High Equivalence only if both a FHIR R4 Core path and, where coded values are required, an internationally-released terminology code were available without modification; profile-level conformance to JP-Core was permitted at this tier provided no JP-Core extension was invoked. Partial Equivalence applied when mapping was achievable without modification to FHIR R4 Core itself, via any of the four mechanisms enumerated in Methods Section 2.2 (JP-Core extension; value-set translation; FHIR Core extension; or Core path/resource structure). Low/No Equivalence applied only when no internationally-recognized semantic counterpart existed and the gap was traceable to a Japan-specific regulatory or administrative framework. Table 1 summarizes the adjudication criteria; the per-entry mechanism applied for each Partial mapping is documented in the JP-Core column of Supplementary Table S1.

**Table 1.** Equivalence Adjudication Criteria.

| Tier | FHIR R4 Core path | Terminology binding | Required modification |
| --- | --- | --- | --- |
| High | Available without modification | LOINC or SNOMED CT international code exists | None |
| Partial | Available without modification to FHIR R4 Core, via JP-Core extension, value-set translation, FHIR Core extension, or Core path/structural use | (a) JP-Core extension; or (b) value-set translation; or (c) FHIR Core extension; or (d) Core path/resource structure | None to FHIR R4 Core itself |
| Low/No | Available as resource, but no terminology binding exists | No internationally recognized code exists; gap is institutional | Requires new terminology submission or cross-mapping table (policy task) |

A large language model (Anthropic Claude) was used as a methodological assistant during the adjudication process to (a) cross-check candidate FHIR R4 paths and LOINC/SNOMED CT codes against the public specifications, (b) generate first-draft equivalence rationales for subsequent human review, and (c) flag candidate inconsistencies in the mapping table for re-evaluation. All terminology look-ups were verified by the lead author against primary specifications (FHIR R4 release, LOINC v2.76, SNOMED CT International browser, JP-Core IG v1.1.1) before inclusion. The model was not used to generate scientific claims, and every equivalence classification, mechanism assignment, and institutional-gap interpretation was finalized by the lead author. Use of AI assistance is further declared in the Author Contributions section.

### 2.4 Pseudonymization Architecture Analysis

We characterized JIPAD’s anonymization architecture from primary documentation to assess its compatibility with FHIR-based record linkage. Specifically, we examined the distinction between pseudonymization (仮名加工化) — which preserves linkage capability — and complete anonymization, as defined under Japan’s Act on the Protection of Personal Information (APPI) 2022 amendments.

## 3. Results

### 3.1 Overall Mapping Equivalence

Of 114 mapping entries representing the 122 JIPAD variables, 97 (85.1%) achieved High Equivalence via direct LOINC or SNOMED CT binding within FHIR R4 Core, and 12 (10.5%) achieved Partial Equivalence via the four mechanisms enumerated in Methods Section 2.2 (JP-Core extension, value-set translation, FHIR Core extension, or Core path/resource structure). Combined technical feasibility reached 95.6% (109/114). Only 5 entries (4.4%) were classified as Low/No Equivalence. Table 2 summarizes the distribution; the complete variable-level mapping with FHIR paths, terminology codes, equivalence rationale, and per-entry mechanism for all 114 entries is provided in Supplementary Table S1.

**Table 2.** Semantic Equivalence Distribution of JIPAD Mapping Entries (n=114)

| Equivalence Category | n | % | Primary Terminology |
| --- | --- | --- | --- |
| High Equivalence (direct FHIR R4 mapping) | 97 | 85.1% | LOINC / SNOMED CT |
| Partial Equivalence (via JP-Core extensions) | 12 | 10.5% | JP-Core IG v1.1.1 |
| Low / No Equivalence (institutional barrier) | 5 | 4.4% | No international standard |
| Total | 114 | 100.0% | — |
| Technical Feasibility (High + Partial) | 109 | 95.6% | Primary finding |

### 3.2 Identified Institutional Gaps

The five Low/No Equivalence variables are detailed in Table 3. All five share a common etiology: Japan’s proprietary disease classification system, which was developed in parallel with — and deeply embedded within — the DPC reimbursement framework, without alignment to ICD-10 or SNOMED CT. This system comprises 288 adult disease codes and 165 pediatric codes, none of which possess established international semantic counterparts.

**Table 3.** Identified Institutional Gaps — Low/No Equivalence Variables.

| JIPAD Variable | FHIR Resource | Equivalence | Barrier ("Wall") | Policy Resolution |
| --- | --- | --- | --- | --- |
| Primary Disease Code (Adult) [Data No. 22] | Condition.code | Low | Wall 1: DPC-specific coding (288 proprietary codes, no ICD-10/SNOMED CT mapping) | Cross-mapping table to ICD-10/SNOMED CT (JSICM × MHLW joint initiative) |
| Secondary Disease Codes ×5 [Data No. 23–27] | Condition.code | Low | Wall 1: Same DPC-specific system (multi-entry, same barrier) | Shared mapping table with primary code |
| Pediatric Primary Disease Code [Data No. 112] | Condition.code | Low | Wall 1: JIPAD-proprietary pediatric codes (165 codes, no ICD-10-CM/PCS mapping) | ICD-10-CM/PCS + SNOMED CT correspondence table (PICU Committee initiative) |
| Pediatric Secondary Codes ×5 [Data No. 113–117] | Condition.code | Low | Wall 1: Same pediatric coding system | Shared with pediatric primary code |
| PMX (Endotoxin Adsorption) [Data No. 42] | Procedure | Low | Wall 4: Japan-specific procedure; no SNOMED CT concept exists internationally | Submit new SNOMED CT concept request via JSICM to SNOMED International |

The sole non-disease-code gap is PMX (polymyxin B-immobilized fiber column hemoperfusion for endotoxin adsorption), a procedure developed and widely used in Japan but absent from SNOMED CT’s international release.

### 3.3 Pseudonymization Architecture

JIPAD employs pseudonymization with record-linkage capability (連結可能匿名化), as explicitly documented in its Data Dictionary (v3.7.2, p.14): patient management numbers are pseudonymized with linkage capability at each facility. Patient-identifying fields (patient ID, name, kana name) are explicitly excluded from central server transmission, as specified in the Linkage Items List.

This architecture enables 99% DPC data matching while preserving anonymity, as documented in the Data Dictionary (p.13). Furthermore, a MHLW-funded DPC data linkage clinical research program commenced in FY2025, representing active institutional progress toward resolving the sole remaining barrier. Table 4 summarizes the evidence base for JIPAD’s pseudonymization architecture.

**Table 4.**
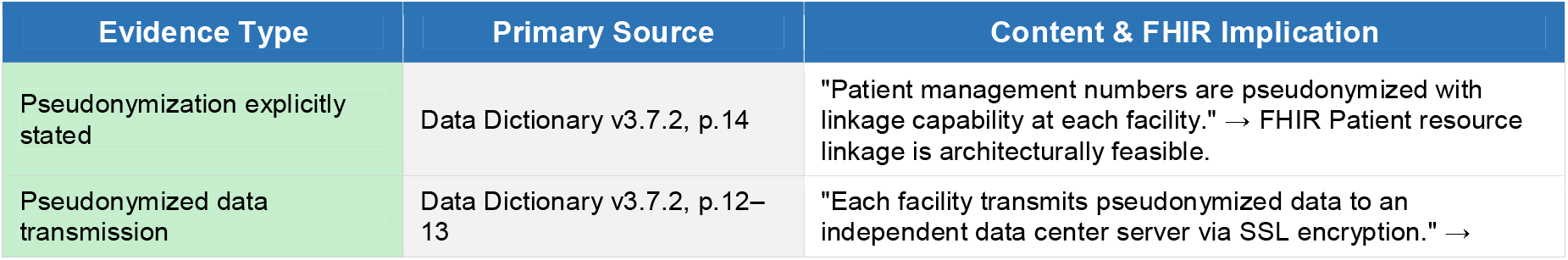
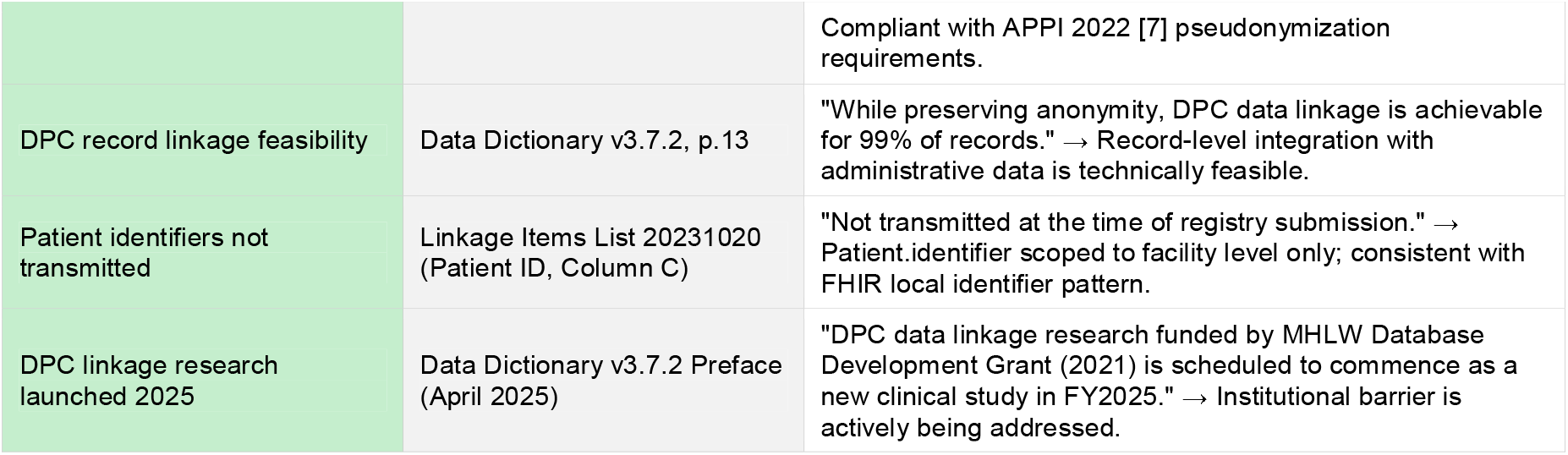
Evidence for JIPAD Pseudonymization Architecture and Record Linkage Capability.

## 4. Discussion

### 4.1 The Inversion of the Expected Problem

The principal finding of this study is an inversion of the expected problem. Prior to this analysis, the assumption — consistent with narrative descriptions of Japan’s health data challenges [1] — was that JIPAD’s isolation reflected technical or design-level incompatibilities with international standards. This assumption is refuted by our data.

JIPAD is technically ready for FHIR integration at a 95.6% level. Its pseudonymization architecture already supports record linkage, and DPC data matching at 99% is documented as feasible. The barriers to integration are neither technical nor architectural. Based on a semi-structured interview with JIPAD’s founding chair (S. Hashimoto, personal communication, March 2025), three interconnected non-technical barriers have been identified:

1. **Organizational barrier:** JIPAD is operated entirely by a volunteer working group (WG) without dedicated funding or staffing for FHIR implementation. This structural under-resourcing means that technically feasible work — for which the data architecture is already prepared — cannot be executed, not due to any design constraint but due to the absence of human and financial capacity.
2. **Epistemic barrier:** JHDS policy stakeholders are currently unaware of JIPAD’s existence, and no formal channel of communication has been established between the JHDS initiative and the JIPAD WG. This mutual invisibility constitutes a structural blind spot in Japan’s health data integration architecture — one that cannot be resolved by technical means alone.
3. **Administrative barrier:** JIPAD integration holds low priority within MHLW’s current policy agenda. This is not a technical judgment but a resource-allocation decision — one that is subject to change if the strategic value of JIPAD data is made visible to policymakers.

This three-barrier framework reframes the problem entirely. The question is not “can JIPAD be integrated into JHDS?” — our data demonstrate it can, at a 95.6% technical feasibility. The question is “who will make it happen, and does the right stakeholder know it needs to happen?” These are organizational and political questions, and they require organizational and political solutions.

### 4.2 Defining “Institutional Debt”

We propose the concept of Institutional Debt to describe systemic, non-technical constraints embedded in healthcare administration that accrue over time and obstruct semantic interoperability. Analogous to technical debt in software engineering — where short-term implementation decisions create long-term maintenance costs — institutional debt arises when administrative systems are built for local operational purposes without anticipating future integration requirements.

Japan’s DPC classification system exemplifies institutional debt. Developed to rationalize hospital reimbursement, it has accumulated 453 proprietary codes (288 adult, 165 pediatric) over two decades, none mapped to ICD-10 or SNOMED CT. The cost of this debt is now externalized onto every downstream integration effort, including JIPAD’s FHIR mapping.

### 4.3 JIPAD as Indispensable Anchor Data for JHDS

A legitimate question arises: given JIPAD’s focus on critical care — a relatively small patient population — what is its strategic importance to the JHDS?

The answer lies in the disproportionate resource concentration of critical illness. ICU patients represent a small fraction of hospitalizations but account for a substantially higher proportion of healthcare expenditure, complex intervention records, and mortality risk — a pattern reflected in Japan’s DPC cost-weight data, where ICU-level episodes consistently carry among the highest per-diem weights in the reimbursement schedule. A health data space that lacks high-acuity critical care data is, by design, blind to the most resource-intensive and clinically consequential episodes of care.

Moreover, JIPAD data is structurally distinct from general electronic health records: it captures time-stamped severity scores, multi-organ support interventions, and granular physiological parameters unavailable in administrative or ambulatory records. These variables are precisely what machine learning-based clinical decision support and outcomes research require.

JIPAD thus serves as anchor data — the high-acuity clinical substrate that gives the JHDS its clinical validity. Without it, the JHDS risks becoming a comprehensive registry of the relatively healthy.

### 4.4 Future Research Directions Enabled by FHIR Integration

FHIR integration of JIPAD would unlock two categories of research currently inaccessible due to data isolation: pharmacoepidemiological analysis and social epidemiological investigation of regional disparities.

First, JIPAD already collects catecholamine administration data for four agents (norepinephrine, dobutamine, dopamine, and epinephrine) across 151 facilities. Once linked to DPC administrative records — a linkage confirmed as 99% feasible by the data architecture documented in this study, and formally initiated as a clinical research program in FY2025 — propensity-score-based pharmacoepidemiological analyses of vasopressor use patterns and their relationship to ICU outcomes will become possible. Such analyses would constitute the first national-level evidence on catecholamine practice variation in Japanese critical care, with direct implications for clinical guideline development.

Second, integration of JIPAD with secondary medical zone infrastructure data (ICU bed density, facility concentration indices) would enable multi-level analysis of regional disparities in critical care outcomes. Japan’s 330 secondary medical zones exhibit substantial variation in ICU resource availability. Whether this structural variation independently predicts patient-level outcomes — after adjustment for severity — is a question that cannot be answered with either dataset alone. FHIR-enabled JIPAD, linked to regional health infrastructure data, would provide the methodological foundation for this inquiry, with direct relevance to Japan’s Regional Healthcare Vision (地域医療構想) policy framework.

Both research directions are technically feasible given the pseudonymization architecture and DPC linkage capability documented in this study. The barrier in each case is not methodological but institutional: the absence of a formal data governance framework permitting cross-registry linkage at scale. FHIR standardization of JIPAD is a necessary — though not sufficient — precondition for realizing this research potential.

### 4.5 Pathway to 2027 JHDS Integration

Recent FHIR implementation initiatives across Asia provide instructive comparators. Indonesia’s national Satusehat platform has implemented an FHIR-based primary care interoperability layer, with documented pain points clustering around server architecture, profile selection, and terminology mapping — the last being directly analogous to the JP-Core extension challenges identified in our analysis [15]. Malaysia’s four-decade trajectory of healthcare digitalisation policy [16] further illustrates that the principal determinants of national-scale interoperability outcomes are governance and policy alignment rather than technical capacity per se. Japan’s position is distinctive in that the technical readiness of its largest critical care registry has already been demonstrated (this study); the remaining gap is exclusively institutional. Against this comparative backdrop, we propose a three-stage policy roadmap for JIPAD-JHDS integration:

Based on our analysis, we propose the following operational steps:

- Near-term (by end of FY2026): Establish a formal JHDS-JIPAD liaison channel — the epistemic barrier must be resolved before any technical work can be commissioned. This requires a single institutional introduction between the JHDS secretariat and the JIPAD WG chair. Concurrently, formalize the MHLW-funded DPC linkage clinical research program and establish a JSICM-MHLW working group to produce an ICD-10/SNOMED CT cross-mapping table for all 288 adult and 165 pediatric JIPAD disease codes.
- Mid-term (FY2027): Pilot FHIR R4-compliant JIPAD data export in 10 volunteer facilities. Validate JP-Core extension profiles against real JIPAD data. Submit a SNOMED CT international concept request for PMX via JSICM.
- Long-term (by 2027): Complete JIPAD-JHDS integration under the national FHIR infrastructure, with JIPAD serving as the critical care data node of the JHDS.

## 5. Limitations

Several limitations warrant acknowledgment. First, the semantic equivalence assessments were performed by a single analyst (K. Ohno); no independent inter-rater reliability assessment was conducted. To partially mitigate this, the adjudication procedure was specified in advance as a fixed, four-stage decision sequence applied uniformly across all 114 entries (Methods Section 2.3, Table 1), and each Partial Equivalence assignment was annotated with the specific mechanism invoked (JP-Core extension, value-set translation, FHIR Core extension, or Core path/resource structure) in the JP-Core column of Supplementary Table S1, allowing independent reviewers to retrace and challenge individual assignments. Nevertheless, all equivalence classifications should be interpreted with this single-analyst caveat in mind, and formal inter-rater validation — including kappa statistics across multiple reviewers with FHIR terminology expertise — is planned as a next step prior to implementation. Second, this mapping is scoped to JIPAD version 3.7.2; future JIPAD versions (4.0 is under discussion) may introduce new variables requiring re-evaluation. Third, while we document JIPAD’s pseudonymization architecture from primary sources, the operational details of the FY2025 DPC linkage research program were not available at the time of writing. Fourth, generalizability of findings to other Japanese clinical registries requires independent evaluation. Fifth, this study presents a semantic mapping framework and gap analysis; it does not include a technical implementation or system validation. The complete variable-level mapping is provided in Supplementary Table S1. The proposed FHIR output pipeline and pharmacoepidemiological and social epidemiological analyses described in Section 4.4 remain as future work, contingent on DPC data linkage completion and institutional governance approval.

## 6. Conclusion

This study demonstrates that JIPAD — Japan’s national ICU registry — is technically and architecturally ready for HL7 FHIR R4 integration at a 95.6% level. The pseudonymization architecture already supports record linkage, and DPC data matching at 99% is feasible.

The remaining 4.4% barrier is exclusively institutional: it lies in MHLW’s governance of Japan’s DPC disease classification system [6], a form of Institutional Debt accumulated over two decades of reimbursement-driven data design. Beyond this technical residue, three non-technical barriers explain why integration has not yet occurred: the JIPAD WG operates on volunteer effort without dedicated resources; JHDS stakeholders are currently unaware of JIPAD’s existence; and MHLW’s current policy agenda assigns this integration low priority. Each of these barriers is resolvable — none requires new technology.

As the nation’s sole high-acuity clinical registry, JIPAD provides anchor data that is structurally irreplaceable in the JHDS. Its integration is not merely beneficial — it is a prerequisite for a clinically valid Japanese Health Data Space by 2027. The path forward is clear. The decision to walk it is institutional.

## Supporting information

Supplemental Table S1. JIPAD to FHIR R4 mapping (xlsx)

## Data Availability

All data supporting this study are publicly available. The complete variable-level mapping is provided in Supplementary Table S1 submitted with this manuscript. Primary sources (JIPAD Data Dictionary v3.7.2, HL7 FHIR R4 specification, JP-Core Implementation Guide v1.1.1, LOINC, SNOMED CT) are publicly accessible at the URLs listed in the References. No patient data were used in this study.

## Author Contributions

K. Ohno: conceptualization, data curation, formal analysis (semantic mapping), methodology, writing — original draft, writing — review and editing. S. Hashimoto: conceptualization, resources (JIPAD primary documentation access), validation, writing — review and editing. Both authors approved the final manuscript and accept full responsibility for its content. Generative AI assistance: A large language model (Anthropic Claude) was used by the lead author (K. Ohno) for first-draft text generation in the Methods and Discussion sections, structural editing of the manuscript, and preliminary cross-checking of FHIR R4 / terminology mappings against public specifications, as detailed in Methods Section 2.3. The AI tool was not used to generate scientific claims, conduct adjudication, or interpret institutional context; the AI tool is not listed as an author and bears no authorial responsibility. All AI-supported content was reviewed, verified against primary sources, and finalized by the human authors, who take full responsibility for the integrity of the work.

## References

[1] Yamamoto I. 医療データ後進国ニッポン、2027 年に間に合うか？『日版 EHDS』10 の壁全解説 [Japan’s lagging health data landscape: will it be ready by 2027? A complete guide to the “Ten Walls” of the Japanese EHDS] [Internet]. note; 2026 [cited 2026 Jul 6]. Available from: https://note.com/kirik/n/n8f9380d879bb. Japanese. [Web commentary by a senior researcher of the Japan Institute of Law and Information Systems (JILIS); cited solely as the conceptual source of the “Ten Walls” typology; not peer-reviewed.]

[2] Benson T, Grieve G. Principles of Health Interoperability: FHIR, HL7 and DICOM. 4th ed. London: Springer; 2021. doi:10.1007/978-3-030-56883-2

[3] HL7 Japan. HL7 FHIR JP Core Implementation Guide v1.1.1 [Internet]. Tokyo: HL7 Japan; 2022 [cited 2026 Apr]. Available from: https://jpfhir.jp/fhir/core/

[4] JSICM ICU Function Evaluation Committee; JIPAD Working Group. JIPAD Data Dictionary Version 3.7.2 [Internet]. Tokyo: Japanese Society of Intensive Care Medicine; 2025 Sep 30 [cited 2026 Apr]. Available from: https://www.jipad.org/images/data/manual/dic_ver3-7-2_20250930.pdf

[5] Yoshihara H. Millennial Medical Record Project: toward establishment of authentic Japanese version EHR and secondary use of medical data (千年カルテプロジェクト). Joho Kanri (Journal of Information Processing and Management). 2018;60(11):767–78. Japanese.

[6] Ministry of Health, Labour and Welfare (Japan). DPC/PDPS: Diagnosis Procedure Combination / Per-Diem Payment System — Data Specifications and Disease Classification Manual [Internet]. Tokyo: MHLW; 2024 [cited 2026 Apr]. Available from: https://www.mhlw.go.jp/stf/seisakunitsuite/bunya/0000049343.html

[7] Act on the Protection of Personal Information (APPI), Act No. 57 of 2003, as amended by Act No. 37 of 2022 (effective April 2022). Government of Japan. Available from: https://www.ppc.go.jp/en/legal/

[8] ISO. ISO 21564:2019 Health informatics — Terminological resources — Requirements for semantic interoperability. Geneva: International Organization for Standardization; 2019.

[9] Regenstrief Institute. LOINC: Logical Observation Identifiers Names and Codes [Internet]. Version 2.76. Indianapolis: Regenstrief Institute; 2024 [cited 2026 Apr]. Available from: https://loinc.org/

[10] SNOMED International. SNOMED CT Browser [Internet]. London: SNOMED International; 2024 [cited 2026 Apr]. Available from: https://browser.ihtsdotools.org/

[11] HL7 International. HL7 FHIR Release 4 (R4) [Internet]. Ann Arbor: Health Level Seven International; 2019 [cited 2026 Apr]. Available from: https://hl7.org/fhir/R4/

[12] Hashimoto S. Semi-structured interview regarding barriers to JIPAD-JHDS integration [personal communication]. 2025 Mar. [Role: JIPAD founding chair; Chair, ICON (Intensive Care Collaboration Network); Professor Emeritus, Kyoto Prefectural University of Medicine.]

[13] European Parliament and Council of the European Union. Regulation (EU) 2025/327 of 11 February 2025 on the European Health Data Space. OJ L, 2025/327, 5 March 2025 (entered into force 26 March 2025). Available from: https://eur-lex.europa.eu/eli/reg/2025/327/oj

[14] Ministry of Health, Labour and Welfare (Japan). Data Health Reform: Vision for the Japanese Health Data Space (JHDS) [Internet]. Tokyo: MHLW; 2023 [cited 2026 Apr]. Available from: https://www.mhlw.go.jp/stf/seisakunitsuite/bunya/kenkou_iryou/iryou/johoka/

[15] Heryawan L, Mori Y, Yamamoto G, Kume N, Lazuardi L, Fuad A, Kuroda T. Fast Healthcare Interoperability Resources (FHIR)-Based Interoperability Design in Indonesia: Content Analysis of Developer Hub’s Social Networking Service. JMIR Form Res. 2025;9:e51270. doi:10.2196/51270

[16] Naim N, Kuroda Y, Yamada S, Mori Y, Liu C, Espinoza R, Yamamoto G, Kuroda T. The evolution of healthcare digitalisation policies in Malaysia: A four-decade narrative review (1985-2025). Digit Health. 2025;11:20552076251353279. doi:10.1177/20552076251353279

